# Are there mortality improvements with newer interventions in adult cardiac surgery? Evidence from 73 meta-analyses

**DOI:** 10.1101/2024.10.31.24316530

**Authors:** Austin Parish, George Tolis, John P.A. Ioannidis

## Abstract

**Background:** In the last two decades, many new interventions have been introduced with the ultimate goal of improving overall postoperative outcomes after cardiac operations in adults. We aimed to assess how often randomized controlled trials (RCTs) in adult cardiac surgery found significant mortality benefits for newer interventions versus older ones, whether observed treatment effect estimates changed over time and whether RCTs and non-randomized observational studies gave similar results.

**Methods:** We searched journals likely to publish systematic reviews on adult cardiac surgery for meta-analyses of mortality outcomes and that included at least one RCT, with or without observational studies. Relative treatment effect sizes were evaluated overall, over time, and per study design.

**Results:** 73 meta-analysis comparisons (824 study outcomes on mortality, 519 from RCTs, 305 from observational studies) were eligible. The median mortality effect size was 1.00, IQR 0.54-1.30 (1.00 among RCTs, 0.91 among observational studies, p=0.039). 4 RCTs and 6 observational studies reached p<0.005 favoring newer interventions. 2/73 meta-analyses reached p<0.005 favoring the newer interventions. Effect size for experimental interventions relative to controls did not change over time overall (p=0.64) or for RCTs (p=0.30), and there was a trend for increase in observational studies (p=0.027). In 34 meta-analyses with both RCTs (n=95) and observational studies (n=305), the median relative summary effect (summary effect in observational studies divided by summary effect in RCTs) was 0.87 (IQR, 0.55-1.29); meta-analysis of the relative summary effects yielded a summary of 0.93 (95% CI, 0.74-1.18).

**Conclusions:** The vast majority of newer interventions had no mortality differences over older ones both overall and in RCTs in particular, while benefits for newer interventions were reported more frequently in observational studies.

## INTRODUCTION

Although adult cardiac surgery was officially introduced as a subspecialty of thoracic surgery 100 years ago with the first closed mitral commissurotomy (1), it did not become widely clinically applicable until more than three decades later, with the development of cardiopulmonary bypass (2). The expanded treatment options afforded by cardiopulmonary bypass transformed the initial scattered heroic interventions by a handful of surgeons into a surgical discipline with a strong clinical and research record. Adult cardiac surgery has been shown to offer very effective interventions for common conditions such as coronary artery disease, various valvulopathies and diseases of the great vessels (3–6), and for some less common ones, e.g. intracardiac and thoracic tumors or abdominal tumors with vascular extension into cardiac chambers (7,8). Many well-established effective interventions are reproducible by a variety of academic and community-based surgeons. Nevertheless, there is also a large and rapidly growing literature in the field attempting to assess newer interventions or modifications of established interventions. Benefits and advantages could come in different forms. A newer intervention may represent an invasive procedure treating a previously medically observed, non-intervenable condition, or may involve a more invasive/aggressive approach than the older procedure which has historically represented the gold standard. Typically, the goal would be to show that the new intervention affords a mortality benefit in order to justify it. Alternatively, a new intervention may not aim to improve mortality, but may be considered to have less invasiveness (and hence perhaps fewer complications and/or lesser cost), therefore rendering it preferable to the established standard-of-care if non-inferiority regarding mortality could be demonstrated.

The gold standard for testing and evaluating treatment interventions is afforded by randomized controlled trials (RCTs), especially when expected effects are likely to be modest and risk-benefit ratios need careful examination. Moreover, it is widely established that when many RCTs exist, careful systematic review and meta-analysis may help examine the overall evidence to hape guidance for eventual clinical decision-making. Many non-randomized studies are also conducted and published. This is even more true for surgical specialties where RCTs may be difficult to conduct (9–11). These non-randomized (henceforth called observational) studies can also be examined in meta-analyses and may also influence eventual guidance and decision-making. The relative merits and comparative outcomes of observational versus randomized evidence has been heavily contested and debated across diverse clinical areas across medicine. Some meta-research evaluations have combined data from multiple meta-analyses where both RCTs and observational studies are available on the same topic (12–14). Some evaluations suggest that, on average, RCTs and observational studies yield similar estimates of the treatment effect (12). Others have found that observational studies tend to have exaggerated estimates of benefit compared with RCTs (13,14). To our knowledge, no such systematic evaluation has been performed including diverse topics in the field of adult cardiac surgery, a field where many observational studies publish estimates of intervention effectiveness.

Here, we undertook a meta-research evaluation that systematically identified and analyzed meta-analyses of adult cardiac surgery interventions where the outcome was mortality and where at least one RCT was available. We aimed to assess how often RCTs in the field found significant benefits for newer interventions versus older ones or doing nothing; whether treatment effect estimates changed over time in the last several decades; and whether RCTs and observational studies on the same intervention comparison give similar results.

## METHODS

### Search strategy and eligibility criteria

This is a meta-research project (15) reported following the PRISMA 2020 checklist for systematic reviews and meta-analyses (16), adapted to our specific meta-research design.

We considered as eligible all systematic reviews and meta-analyses (SRMAs) related to adult cardiac surgery that contained randomized controlled trials (RCTs) (regardless of whether observational non-randomized studies were also included or not) and summarized all-cause mortality outcomes. We searched in publication venues likely to publish most of these adult cardiac surgery SRMAs. Specifically, the search strategy captured for further screening all SRMAs published in the Journal of Thoracic and Cardiovascular Surgery and Annals of Thoracic Surgery (the two primary journals specializing in the field of interest) and all SRMAs that were likely to address cardiac surgery in any of the 4 highest impact factor surgical journals, the 4 highest impact factor general medical journals, and the comprehensive Cochrane Database of Systematic Reviews. The complete search string was ((Annals of Surgery [SO] OR Surgery [SO] OR JAMA Surgery [SO] OR British Journal of Surgery [SO] OR New England Journal of Medicine [SO] OR JAMA [SO] OR Lancet [SO] OR BMJ [SO] OR Cochrane Database of Systematic Reviews [SO]) AND (’cardiac surgery’ OR ’heart surgery’ OR ’coronary bypass’ OR CABG OR ’valve surgery’)) OR (ann thorac surg[Journal] OR j thorac cardiovasc surg[Journal]) AND (systematic[sb] OR meta-analysis[sb]). The search was last updated on August 1^st^, 2023.

Two authors (GT/AP) screened the abstracts of all retrieved items for eligibility. Items were excluded if they were not SRMAs, not studying the effect of interventions on all-cause mortality, contained no RCTs, or were studying topics unrelated to current adult cardiac surgery. When SRMAs on the same comparison and outcomes (duplicates) were identified, the SRMA with most RCTs was included; with ties, we included the one that had more observational studies.

### Data extraction from eligible meta-analyses

For all included eligible SRMAs, we extracted journal, year of publication, and data for all-cause mortality outcomes, including the specific outcome, effect metric (odds ratio [OR], risk ratio [RR], hazard ratio [HR], or incident rate ratio [IRR]), specific comparison (experimental and control groups), the overall reported meta-analysis summary effect and effects from individual studies (summarized in tables or forest plots). For each individual study, we extracted the effect size, 95% confidence interval (CI), and the 2×2 table whenever available. When SRMAs presented all-cause mortality outcomes for different timepoints, all timepoints were extracted.

The summary estimate presented by the authors in the SRMA was extracted. When both adjusted and unadjusted study results were synthesized, we preferred the latter. For 22 SRMAs and a total of 221 individual studies, a summary estimate was not available, but we could calculate OR from 2×2 tables of the individual studies and obtain the summary OR thereof.

For each comparison, we recorded whether the comparison was between an active experimental group and usual care, placebo (sham) or an active control group. We also assessed and recorded the direction of each eligible comparison. Whenever an older intervention was the experimental group and a newer intervention was the control group, we inverted the effect sizes (e.g. 3 became 1/3=0.33), so that eventually all comparisons in our analysis reflect the comparison of a newer intervention versus an older one (or nothing/placebo/sham). Accordingly, effect sizes (relative risks) <1.0 represent a reduction in mortality favoring the newer intervention. Whether such coining (inversion) was needed was determined by consensus between all authors (AP/GT/JPAI) regarding which intervention is newer.

### Validation of data from primary studies

We randomly selected 30 individual studies from the 34 SRMAs that contained both RCTs and observational studies (from a total of 400 individual studies, n=95 RCTs and n=305 observational studies) and re-calculated the study effect to confirm the numerical effect reported by the SRMA authors.

### Statistical techniques

Random effects meta-analysis was carried out using the Sidik-Jonkman estimator (17). All effects were log-transformed before meta-analysis. Heterogeneity was estimated with the Higgins and Thompson’s I^2^ statistic and Cochran’s Q test (18,19). Meta-analysis calculations were performed using the meta package in R (20). Continuous data was also summarized using medians and interquartile ranges (IQR); the Spearman correlation coefficient was calculated between continuous variables (21).

All comparisons of interventions were classified into two types, type A (comparing a newer less invasive method versus an older more invasive method looking at non-inferiority) and type B (comparing a newer more invasive method versus an older less invasive method looking at superiority). Type B includes also all comparisons against doing nothing (or giving placebo/sham). Sensitivity analyses were performed where the summary effect was inverted for all type A comparisons.

For those SRMAs that included also observational studies, we compared the results of RCTs versus observational studies in each topic by obtaining the relative summary effect, i.e. the fixed effects summary effect across observational studies divided by fixed effects summary effect across RCTs) (22). We then synthesized the relative summary effects across all topics that included the RCTs and observational studies. A relative summary effect <1.0 suggests that the observational studies show better results for the newer intervention regarding mortality than RCTs.

Statistical significance was assessed at two levels, α = 0.05 (suggestive significance) and α = 0.005 (formal statistical significance), as proposed by Benjamin et al. (23).

R version 4.1.0 was used for all calculations (R Core Team 2021).

## RESULTS

### Eligible SRMAs and single studies

Our literature search identified 927 items. Of these, 531 (57%) were unrelated to cardiac surgery, 170 (18%) included no RCTs (18%), 50 (5%) did not include mortality outcomes, 26 (3%) were not SRMAs presenting extractable data, 23 (2%) were not studies of interventions, 31 (3.3%) were repeat topics, and 1 was withdrawn. Of the 95 remaining publications of SRMAs, 61 (64%) were found to have extractable data on mortality from forest plots and were included (**Figure 1**). For the other 34 that had no clearly extractable data per study in forest plots, the corresponding authors were contacted, but none offered usable data. Of the 61 publications, 25 (41%) were published in the Journal of Thoracic and Cardiovascular Surgery, 22 (36%) in Annals of Thoracic Surgery, 12 (20%) in the Cochrane Database of Systematic Reviews, and 2 (3%) in Annals of Surgery.

**Figure 1.**
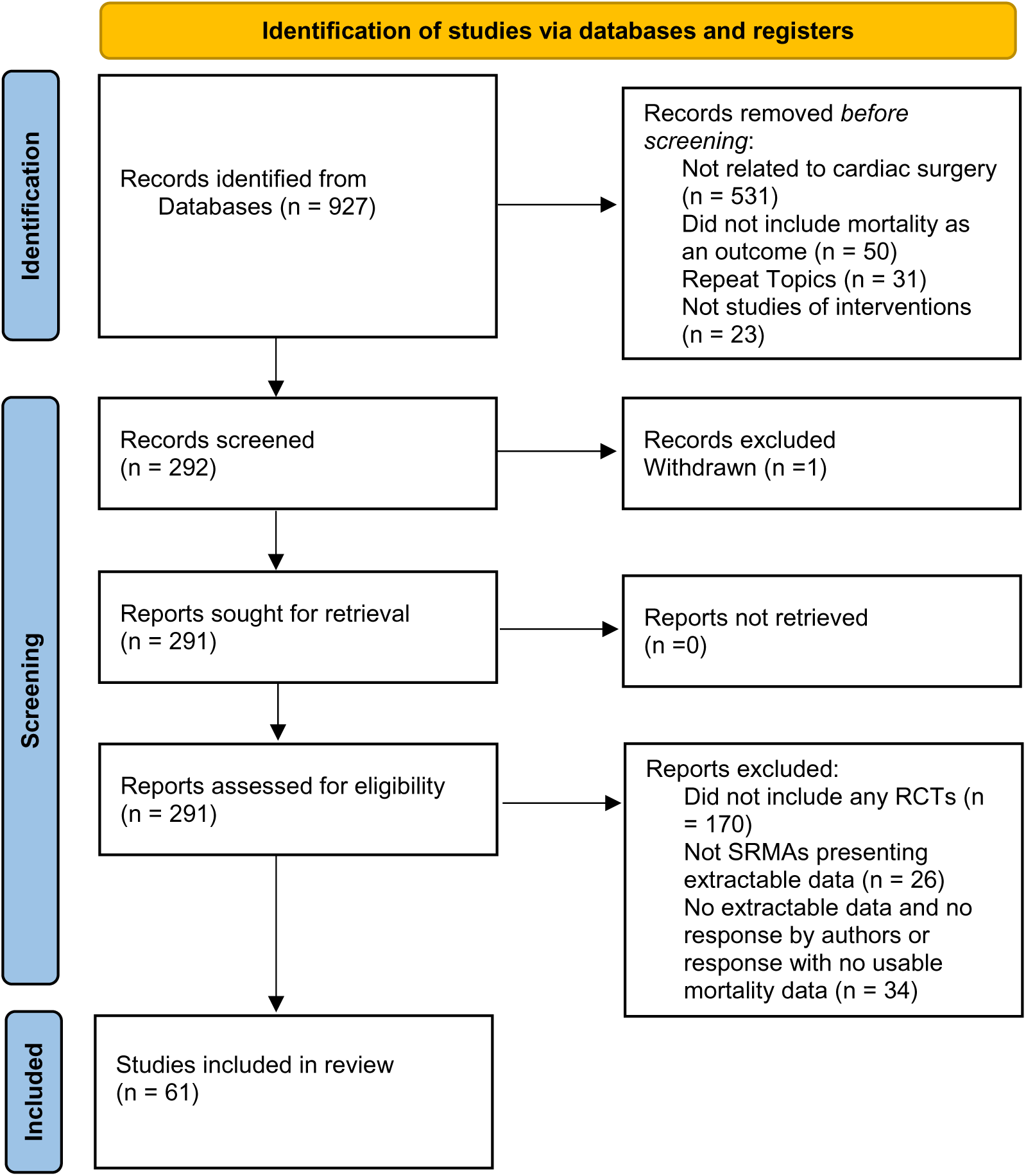
Flow chart for eligible meta-analyses

10 of the 61 papers contained two or more different eligible comparisons (n=2 papers) or timepoints for the comparison of the same intervention (n=8 papers). Overall, 73 different comparisons were eligible for our analyses; of them, 6 (8%) were inverted to consistently have the newer (experimental) intervention compared against the older (control) intervention. The 73 comparisons are shown in **Supplementary Table 1**. They included a total of 824 mortality study outcomes, 519 RCTs and 305 observational ones. Of the 824, 490 were presented as OR, 242 RR, 55 HR, and 37 IRR metrics. Across the 73 meta-analyses, each contained a median of 10 studies (IQR 6 to 17). Of the 73 meta-analyses, 29 (416 study outcomes) were type A comparisons and 44 (408 study outcomes) were type B comparisons.

### Effect sizes and statistical significance in single studies

The median mortality effect size across 824 individual study outcomes was 1.00 (IQR, 0.54-1.30), suggesting no difference in mortality, on average, between experimental and control interventions. Across 519 RCTs, the median effect size was also 1.00 (IQR, 0.58-1.32), whereas across 305 observational studies it was 0.91 (IQR, 0.50-1.26). The difference between RCTs and observational studies suggested statistical significance (Wilcoxon rank sum p=0.039).

Across the 519 RCTs, 36 (6%) had suggestive significance (p<0.05), but only 11 (2%) were formally statistically significant with p=0.005. Suggestive and formal significance was seen in 18 and 4 RCTs in favor of the newer (experimental) intervention and in 18 and 7 RCTs in favor of the control intervention. When limited to the 277 RCTs of type B (presumably superiority) comparisons, 16 (6%) and 5 (2%) had suggestive and formal significance and almost always this favored the newer intervention (**Table 1**).

**Table 1.**
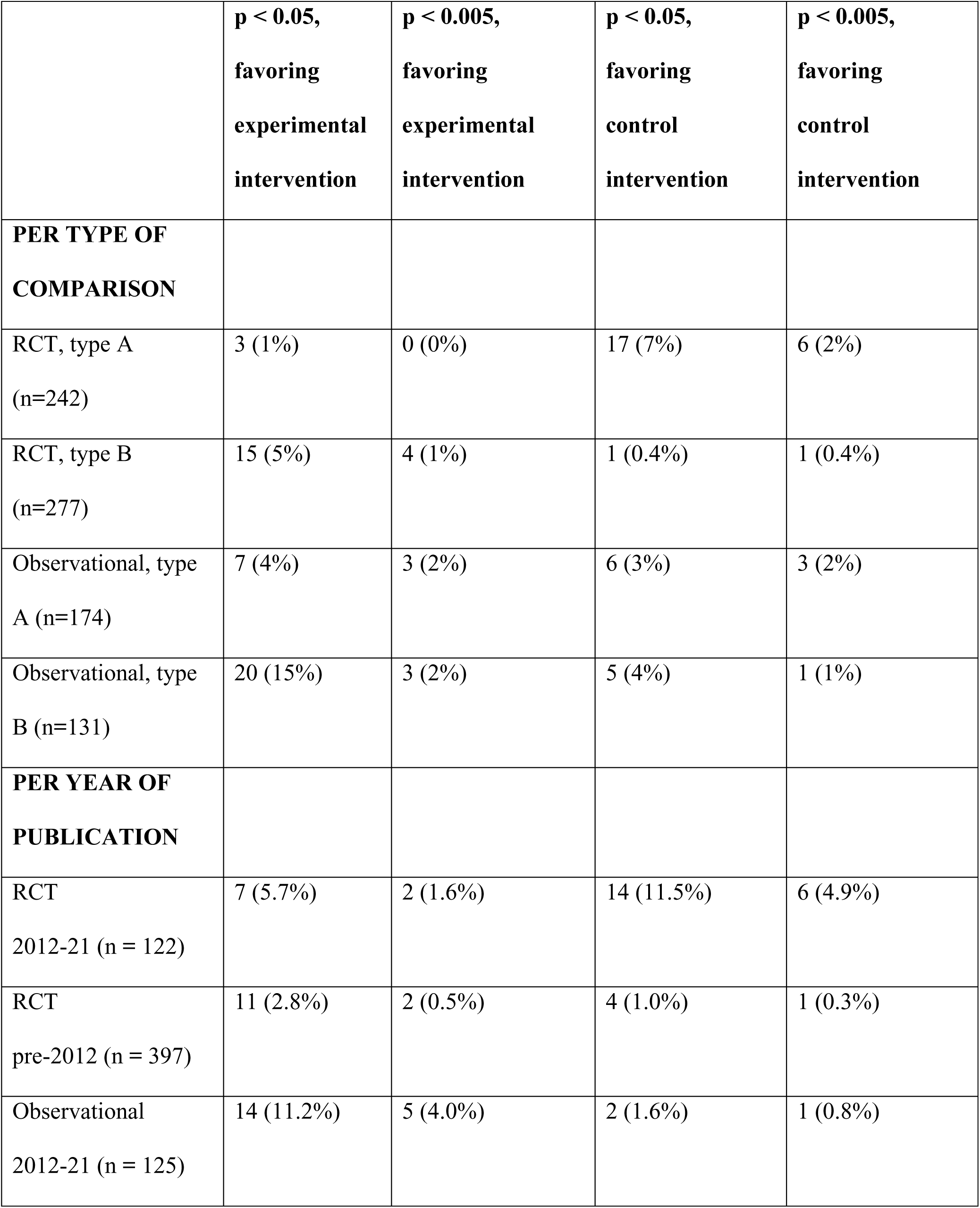
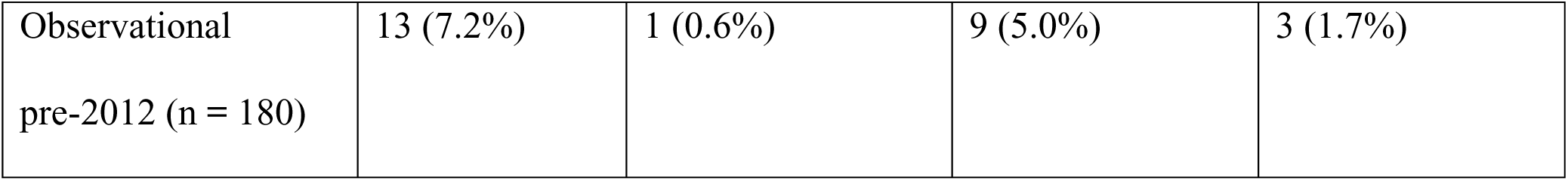
RCTs and non-randomized observational studies with suggestive (p<0.05) and formally statistically significant (p<0.005) results according to type of comparison and year of publication. Type A comparisons involve comparing a newer less invasive method versus an older more invasive method, thus presumably demonstration of non-inferiority might be sufficient to make the newer approach attractive. Type B comparisons involve comparing a newer more invasive method versus an older less invasive method, thus presumably demonstration of superiority would be needed to show that the added invasiveness is worthwhile. Type B includes also all comparisons against doing nothing (or giving placebo/sham).

|  | **p < 0.05,**<br>**favoring**<br>**experimental**<br>**intervention** | **p < 0.005,**<br>**favoring**<br>**experimental**<br>**intervention** | **p < 0.05,**<br>**favoring**<br>**control**<br>**intervention** | **p < 0.005,**<br>**favoring**<br>**control**<br>**intervention** |
| --- | --- | --- | --- | --- |
| **PER TYPE OF**<br>**COMPARISON** |  |  |  |  |
| RCT, type A<br>(n=242) | 3 (1%) | 0 (0%) | 17 (7%) | 6 (2%) |
| RCT, type B<br>(n=277) | 15 (5%) | 4 (1%) | 1 (0.4%) | 1 (0.4%) |
| Observational, type<br>A (n=174) | 7 (4%) | 3 (2%) | 6 (3%) | 3 (2%) |
| Observational, type<br>B (n=131) | 20 (15%) | 3 (2%) | 5 (4%) | 1 (1%) |
| **PER YEAR OF**<br>**PUBLICATION** |  |  |  |  |
| RCT<br>2012-21 (n = 122) | 7 (5.7%) | 2 (1.6%) | 14 (11.5%) | 6 (4.9%) |
| RCT<br>pre-2012 (n = 397) | 11 (2.8%) | 2 (0.5%) | 4 (1.0%) | 1 (0.3%) |
| Observational<br>2012-21 (n = 125) | 14 (11.2%) | 5 (4.0%) | 2 (1.6%) | 1 (0.8%) |
| Observational<br>pre-2012 (n = 180) | 13 (7.2%) | 1 (0.6%) | 9 (5.0%) | 3 (1.7%) |

Across the 305 observational studies, there were 38 (13%) and 10 (3%) with suggestive and formal statistical significance. Most of those favored the newer (experimental) intervention (27 and 6, respectively) and fewer favored the control intervention (11 and 4, respectively). When limited to 131 type B (presumably superiority) comparisons, 25 (19%) and 4 (3%) showed suggestive and formal significance, almost always favoring the newer intervention (**Table 1**).

### Treatment effects over time

A total of 122 RCTs and 125 observational studies had been published in the last decade (2012 or later). The median effect size of these 247 studies was 1.00 (IQR, 0.50-1.26) overall (1.00, IQR, 0.60-1.36 in RCTs and 0.86, IQR, 0.46-1.05 in observational studies). The majority of statistically significant RCTs with p<0.005 (8/11, 73%) were published in the last decade and of these, 6/8 (75%) were in favor of the control intervention. Conversely, among observational studies with p<0.005, 6/10 (60%) had been published in the last decade and 5/6 (83%) were in favor of the newer (experimental) intervention (**Table 1**).

There was no significant change in the effect size for newer versus control interventions over time (p=0.56, p=0.55 after controlling for specific comparisons). For RCTs, there was no significant time effect (p=0.42; after adjusting for specific comparisons, p=0.40). For observational studies, there was a suggestion of a time association with a trend for increasing effects over time (p=0.049; after adjusting for specific comparisons, p=0.065) (**Figure 2**).

**Figure 2.**
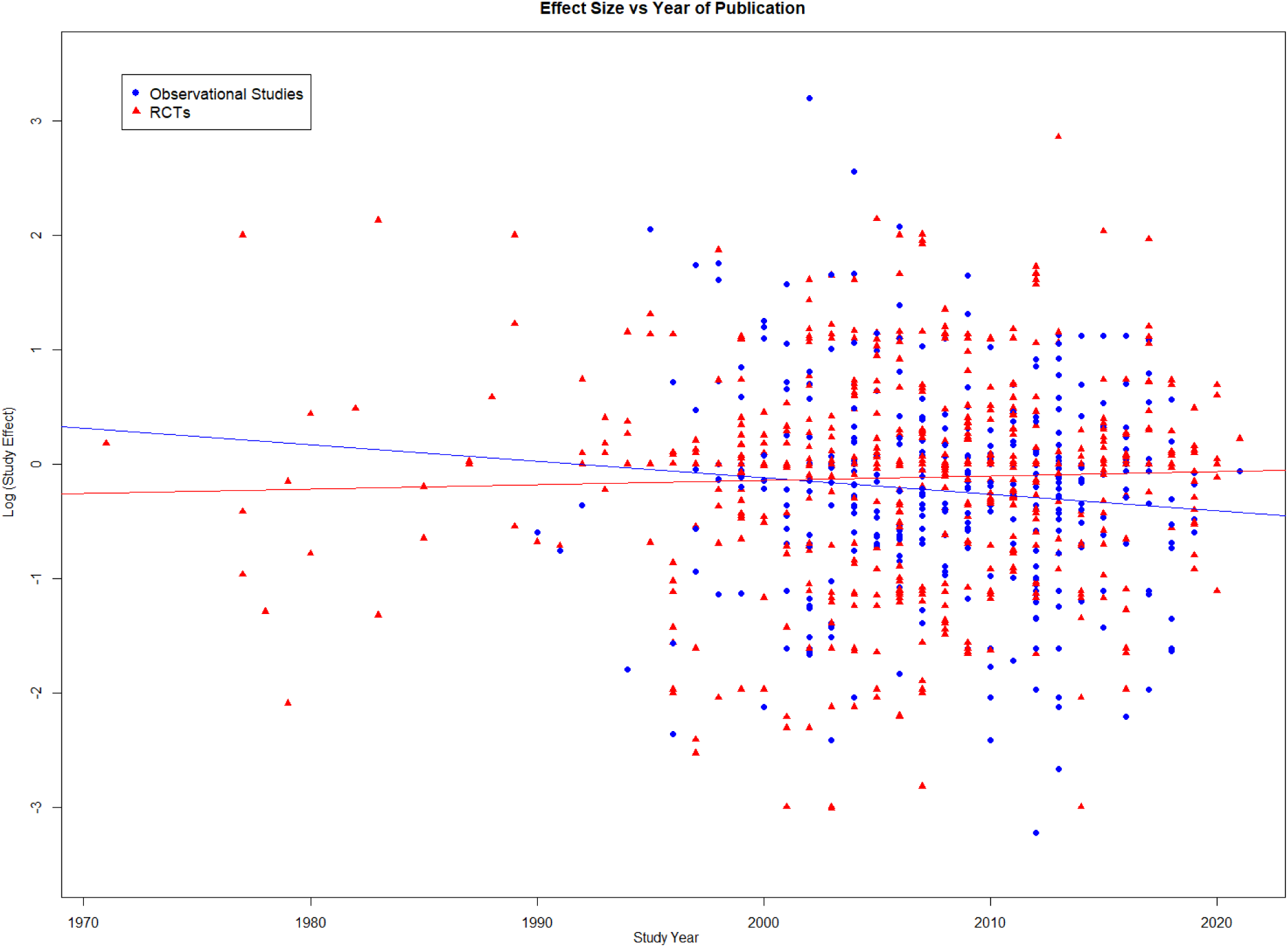
Study effect size vs year of publication, with associated line of best fit, for observational studies (n = 305) and RCTs (n = 519)

### Treatment effects in meta-analyses

As reported by the authors, the median meta-analysis summary effect across 73 comparisons was 0.90 (IQR, 0.69-1.12). Of the 73 meta-analyses, as reported by the authors, 15 (21%) had summary effects with suggestive significance with p<0.05 and 4 (5%) had summary effects that were statistically significant with p<0.005 level in favor of the experimental arm, while 6 (8%) and 3 (4%) meta-analyses had effects at these significance levels favoring the control arm. When we re-analyzed the data with random effects and the SJ method, the respective numbers were 5, 2, 3 and 1.

When limited to data from the 519 RCTs outcomes, the median meta-analysis summary effect was 0.94 (IQR, 0.74 to 1.19); of the 73 meta-analyses based on RCT data, only 2 were statistically significant (p<0.005) in favor of the experimental arm and 2 in favor of the control arm.

When limited to type B (presumably superiority) comparisons (44 meta-analyses total), 14 meta-analyses had summary effects with suggestive significance with p<0.05 and 4 had summary effects that were statistically significant with p<0.005 in favor of the experimental arm; conversely, only 1 and 0 meta-analyses had effects at these significance levels favoring the control arm. When limited to data from the RCTs, the respective numbers were 7, 2, 0, and 0.

### Clinical topics with statistically significant differences

**Table 2** shows the 7 clinical topics where there were statistically significant differences in mortality at the p<0.005 level in the meta-analyses, as originally reported by the systematic review authors, 3 of which retained p<0.005 in our re-calculated SJ random effects estimates.

**Table 2.** Clinical topics with statistically significant survival differences at p<0.005 in meta-analyses AS: aortic stenosis, CABG: Coronary artery bypass graft, CI: confidence interval, IABP: Intra-aortic balloon pump, PCI: percutaneous coronary intervention Follow-up shows the time of mortality assessment.

| Intervention (type A or B) | Condition/ Setting | Metric | Author reported meta-analysis effect | SJ random effects meta-analysis effect | RCTs only random effects meta-analysis effect | Mortality follow-up |
| --- | --- | --- | --- | --- | --- | --- |
| Loading dose vs regular dose of statins (B) | Prophylactic after CABG | OR | 0.74 (0.60-0.91) | 0.78 (0.57-1.06) | 0.78 (0.57-1.06) | 1 to 10 years |
| Off-pump vs on-pump (A) | CABG | HR | 1.07 (1.03-1.11) | 1.04 (0.95-1.14) | 1.20 (0.81-1.76) | 5 year |
| IABP vs No IABP (B) | Pre-op prophylactic before CABG | OR | 0.35 (0.23-0.53) | 0.36 (0.20-0.65) | 0.25 (0.11-0.56) | In-hospital or 30-day |
| Radial artery versus saphenous vein as the second conduit (B) | CABG | IRR | 0.74 (0.63-0.87) | 0.72 (0.54-0.95) | 0.94 (0.61-1.44) | 0.7 to 11.9 years |
| Early surgery vs conservative management (B) | Asymptomatic severe AS | HR | 0.49 (0.36-0.68) | 0.48 (0.32-0.72) | 0.33 (0.12-0.91) | 1.8 to 6.2 years |
| TAVR PCI vs SAVR CABG (A) | CABG | HR | 1.35 (1.11-1.65) | 1.34 (1.05-1.71) | 1.34 (1.05-1.71) | 1 to 3 years |
| PCI vs CABG (A) | Diabetic patients | OR | 1.64 (1.30-2.08) | 1.63 (1.23-2.17) | 1.63 (1.23-2.17) | 5 year |

### Relative effects in RCTs and observational studies

Of 73 meta-analyses, 34 (47%) contained at least one RCT and at least one observational study; across these 34 meta-analyses there were 400 individual studies (95 were RCTs (24%) and 305 observational studies). The median observational summary effect was 0.81 (IQR, 0.68-1.05) vs 0.93 (IQR, 0.77-1.22) in the respective RCTs (paired Wilcoxon p=0.17). The Spearman correlation between RCT and observational summary effect sizes was -0.023 (p=0.90).

The relative summary effect (summary effect in observational studies divided by summary effect in RCTs) had a median of 0.87 (IQR, 0.55-1.29); meta-analysis of the relative summary effects yielded a summary of 0.93 (95% CI 0.74-1.18, p=0.57, I^2^=0%, 0-39%) (**Figure 3**). Respective results were 0.86 (IQR, 0.41-1.04) and 0.85 (95% CI, 0.55-1.34, p=0.85, I^2^= 0%, 0%-57%) for type A comparisons and 1.08 (IQR, 0.78-1.44) and 0.98 (95% CI, 0.75-1.27, p=0.85, I^2^=0%, 0%-47%) for type B comparisons.

**Figure 3.**
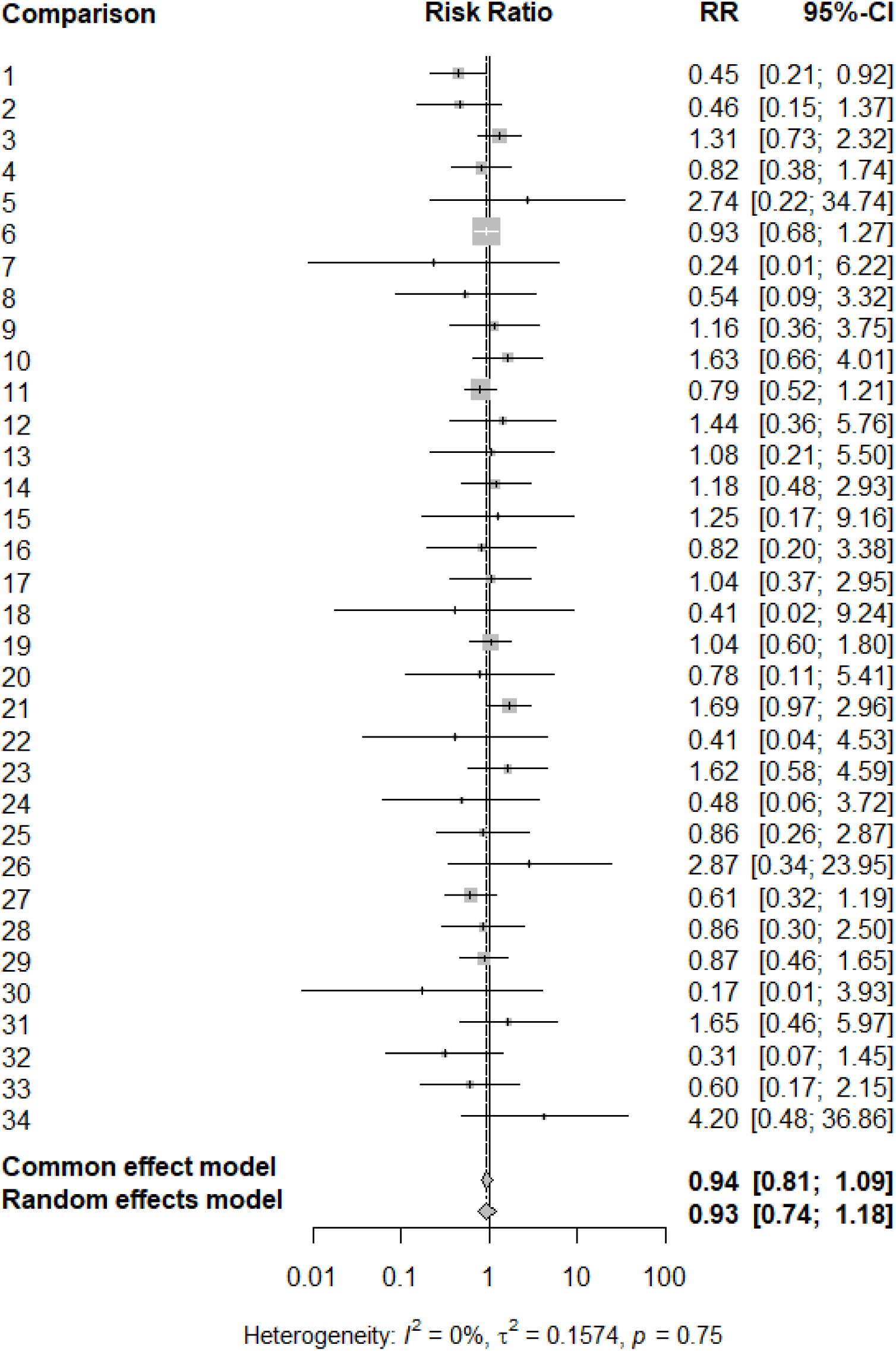
Forest plot of relative summary effect (summary effect in observational studies divided by summary effect in RCTs).

Sensitivity analyses yielded similar results (see **Supplementary Results**).

### Validation of primary data

In the 30 randomly selected studies (from 22 SRMAs), our re-calculated study effect was always the same or very similar (maximum deviation 0.04 in relative risk scale) to the effect reported by the SRMA authors.

## DISCUSSION

Our analysis of 73 comparisons of newer interventions versus control management in adult cardiac surgery shows that mortality benefits of new interventions versus older approaches were uncommon. The median effect size of 1.00 suggests no difference in mortality between the compared groups. The same picture was seen when examining strictly the data from RCTs. Conversely, on average, observational data suggested some benefit for the experimental intervention groups.

Evidence from RCTs rarely suggested statistically significant benefits of the experimental intervention at the level of single studies or meta-analyses thereof. Conversely, a modest fraction of non-randomized observational data suggested significant benefits for the experimental arm. Effect sizes in RCTs have been steady over time, while effect sizes for non-randomized observational data may tend to become larger in more recent years.

Only about half of the 73 topics that we examined included both RCTs and observational data in their meta-analyses. Moreover, very few RCTs were included in these topics where a direct comparison of effect sizes RCTs versus observational studies would be feasible within the same topic. Therefore, the results from these direct comparisons have large CIs and are underpowered to detect clear differences between the two designs. Nevertheless, the picture in this subset is compatible with the overall picture across all 73 comparisons.

There is debate about the merits and uses of non-randomized observational studies to evaluate comparative effectiveness (24,25). Our findings suggest caution in making definitive inferences and formulating guidelines about comparative effectiveness based on observational data in adult cardiac surgery. Observational data have more degrees of freedom and this may result in higher chances of selective reporting that fits some expected narrative. RCTs certainly have their own biases and they are not infallible gold standards. Nevertheless, the effort to promote the use of RCTs in surgery is welcome (26).

The dearth of well-documented benefits in mortality with newer, experimental interventions in adult cardiac surgery does not mean that the field does not make progress. Half of the data that we analyzed pertained to type A comparisons, where the newer intervention was a less invasive one and thus the goal would probably not be to show necessarily better survival, but other comparative benefits, e.g. fewer complications, better convenience or lesser cost. Moreover, among the comparisons that had a superiority outlook with the newer intervention being more aggressive than the control, we could identify two where the newer intervention did significantly better in mortality outcomes than the control at p<0.005. Both of them reflected classic, widely-used interventions where the magnitude of the survival benefit was very large, i.e. use of intra-aortic balloon pump and surgery for asymptomatic severe aortic stenosis. For new and future interventions, one may need to be prepared to detect much smaller but still clinically meaningful survival benefits. This would require larger studies to have sufficient power to detect such benefits with certitude.

We evaluated results both at the level of effect sizes and at the level of statistical significance. Effect sizes are preferable to convey the magnitude of the potential benefit (27). Nevertheless, we should caution that the data are typically presented in relative effect metrics. Clinical decision-making should also examine carefully the absolute magnitude of benefits. Also for statistical significance, some authors argue that dichotomizing p-values may be spurious and misleading and have even urged to abandon the notion of statistical significance (28). Nevertheless, for clinical trials and their SRMAs (as analyzed here), the notion of statistical significance still has relevance: its abandonment may open the door to even more spurious practices, where investigators may move the goalposts without any statistical rules (29). The use of p<0.005 instead of p<0.05 is recommended to avoid false-positive results (23). This suggestion has also empirical grounding in RCTs and meta-analyses thereof (30).

Our study has some limitations. First, we only focused on SRMAs published in the key journals that are likely to publish SRMAs in this specialty, as it would have been inconvenient and low-yield to screen hundreds of thousands of SRMAs published across the entire medical literature. Second, SRMAs may not have been performed yet on some interesting topics and this may affect more prominently recent developments in the field with very recent data. Third, while most of the included data used OR metrics, some used other relative effect metrics and these are not identical. Nevertheless, for the mostly small effects analyzed here, differences between OR and HR, RR, or IRR are probably small. Fourth, while we took meticulous care with data, we depended in existing published SRMAs and these evidence syntheses may include their own biases and errors. There are many efforts to improve the methodological rigor of conduct and the quality of reporting of both primary studies and of SRMAs (31). The importance of enhanced attention to these matters cannot be overstated. Fifth, we could not evaluate whether improved outcomes with new interventions may exist specifically for subgroups of patients, e.g. in recent years there has been an expansion of the pool of surgical candidates. Patients who were considered inoperable in the past due to advanced age or underlying conditions are routinely offered surgery in the current era. Finally, no single study and no single SRMA can be taken as perfect evidence, no matter how well it is done. Therefore, uncertainty about the comparative effects of different interventions may be larger than actually observed.

Allowing for these caveats, our meta-research analysis offers a bird’s eye view of over 800 study effect size estimates from comparisons involving adult cardiac surgery These data can be used as a benchmark for what might be reasonable expectations for future RCT and comparative effectiveness research and SRMAs in the field.

## Data Availability

All data are available from the authors upon request

## Funding

none

## Disclosure statement

The authors have no conflicts of interest.

## Data sharing statement

All data are available from the authors upon request.

Independent data access and analysis: The corresponding author as well as the remaining two authors had full access to all the data in the study. The corresponding author takes responsibility for its integrity and the data analysis.

## Abbreviations

HR: hazard ratio
OR: odds ratio
RCT: randomized controlled trial
RR: risk ratio
SRMA: systematic review and meta-analysis

---

**Supplementary Table 1.**
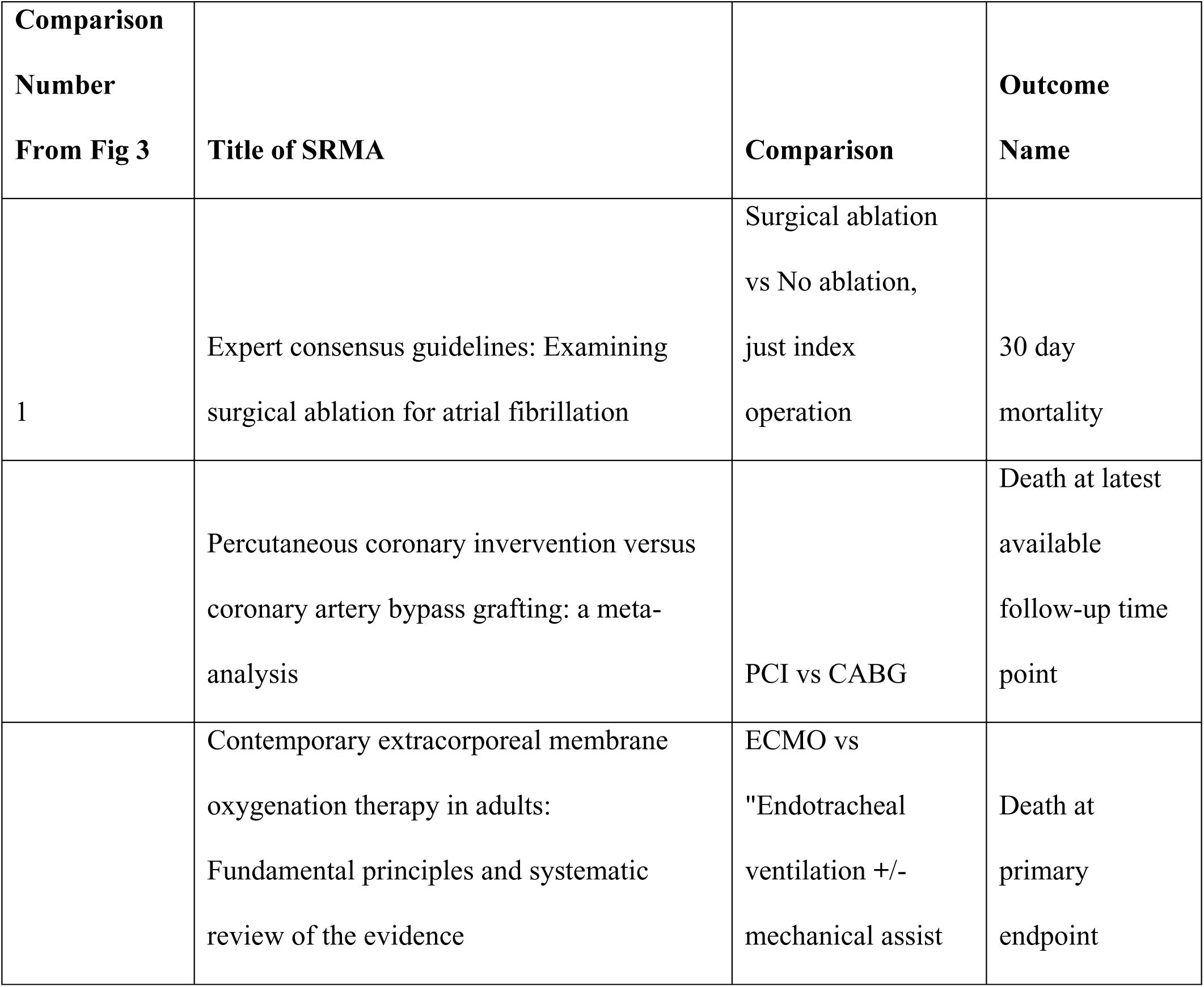
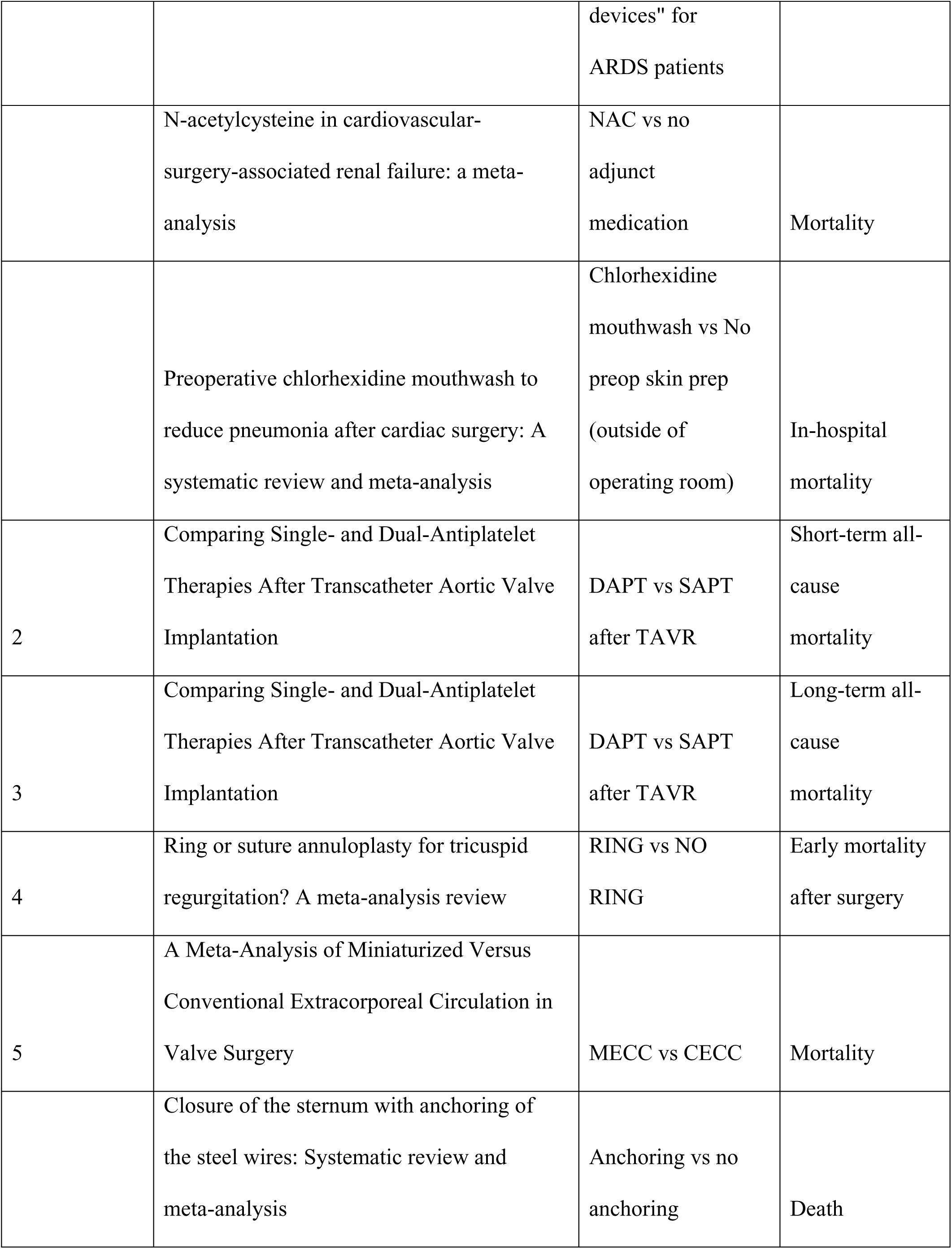
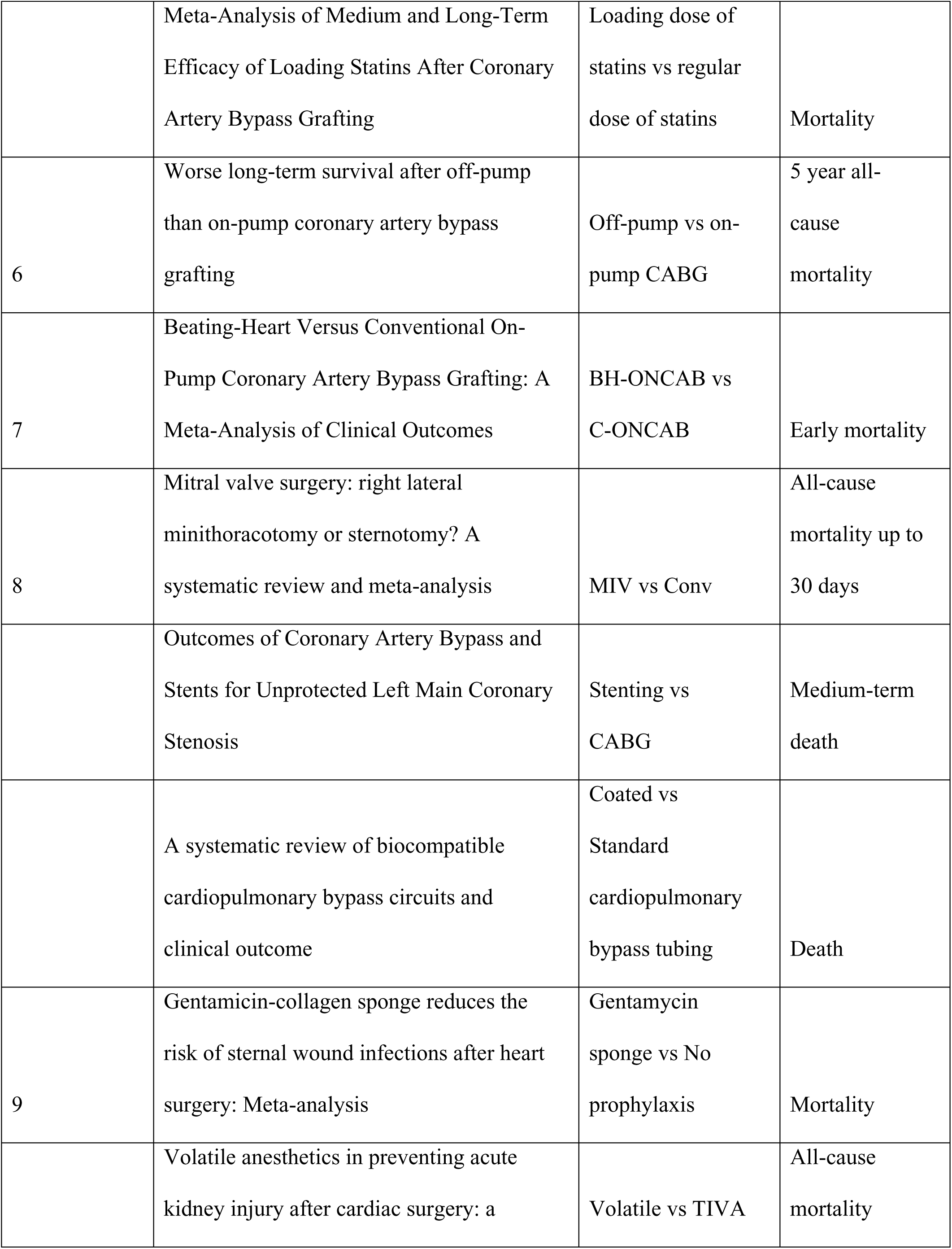
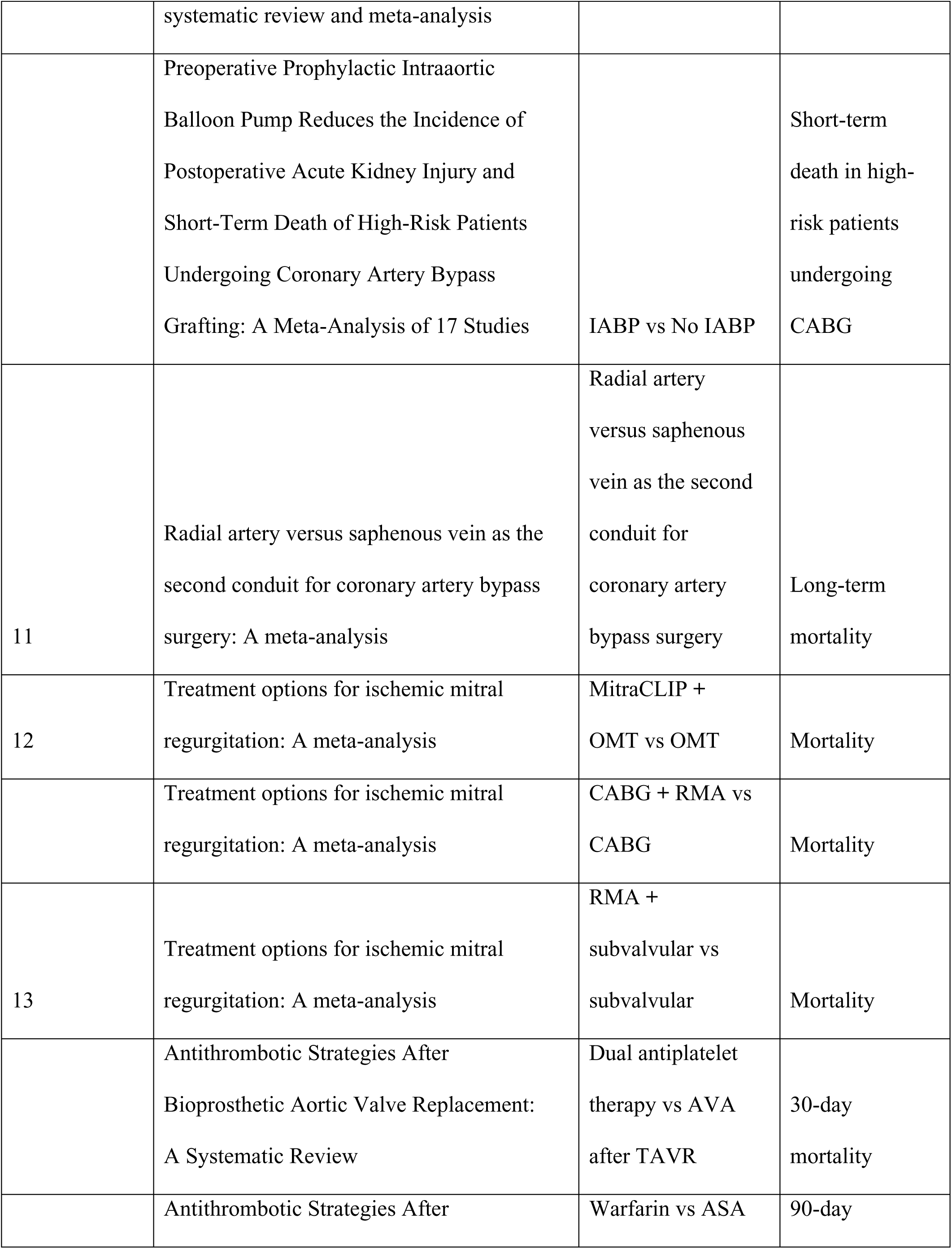
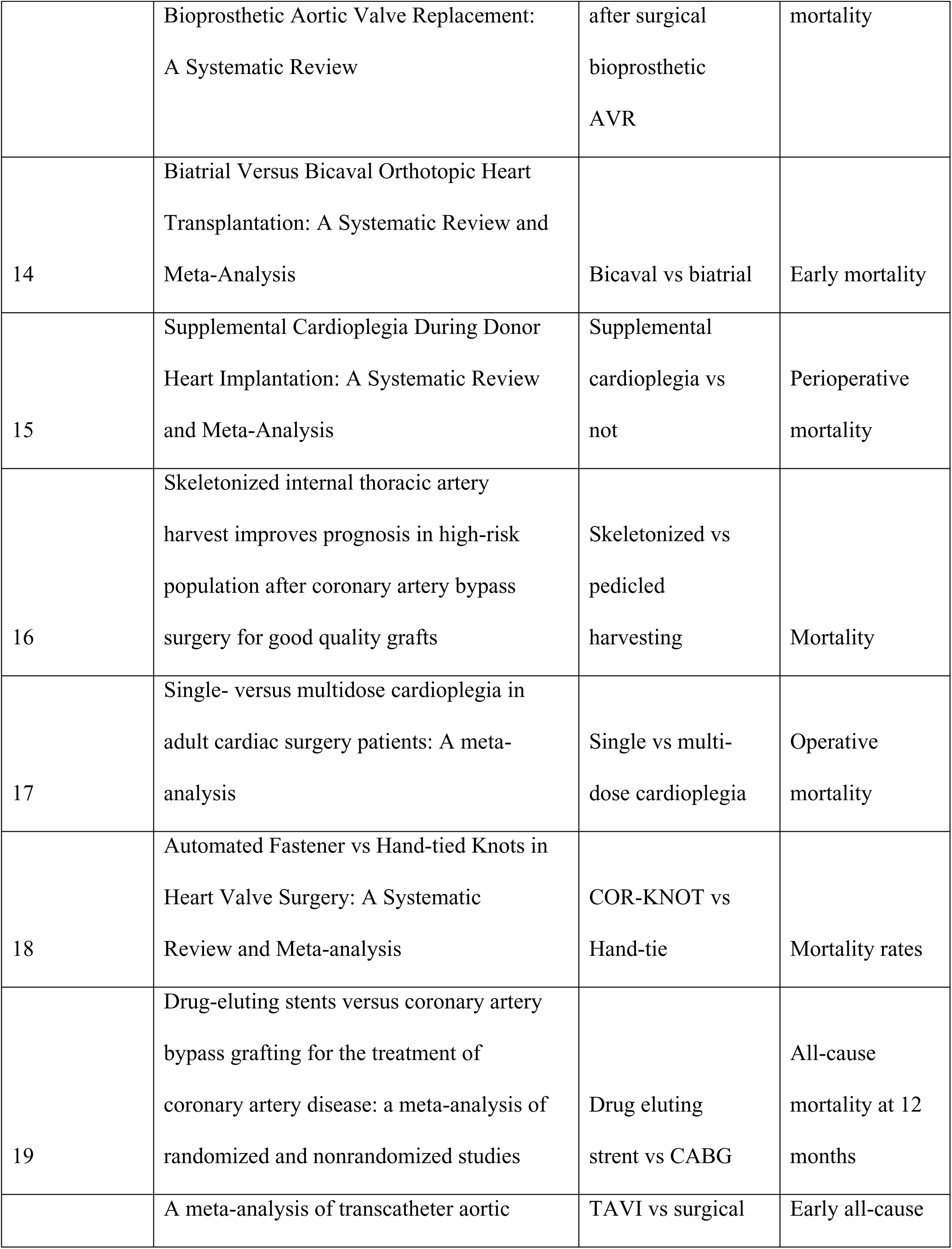
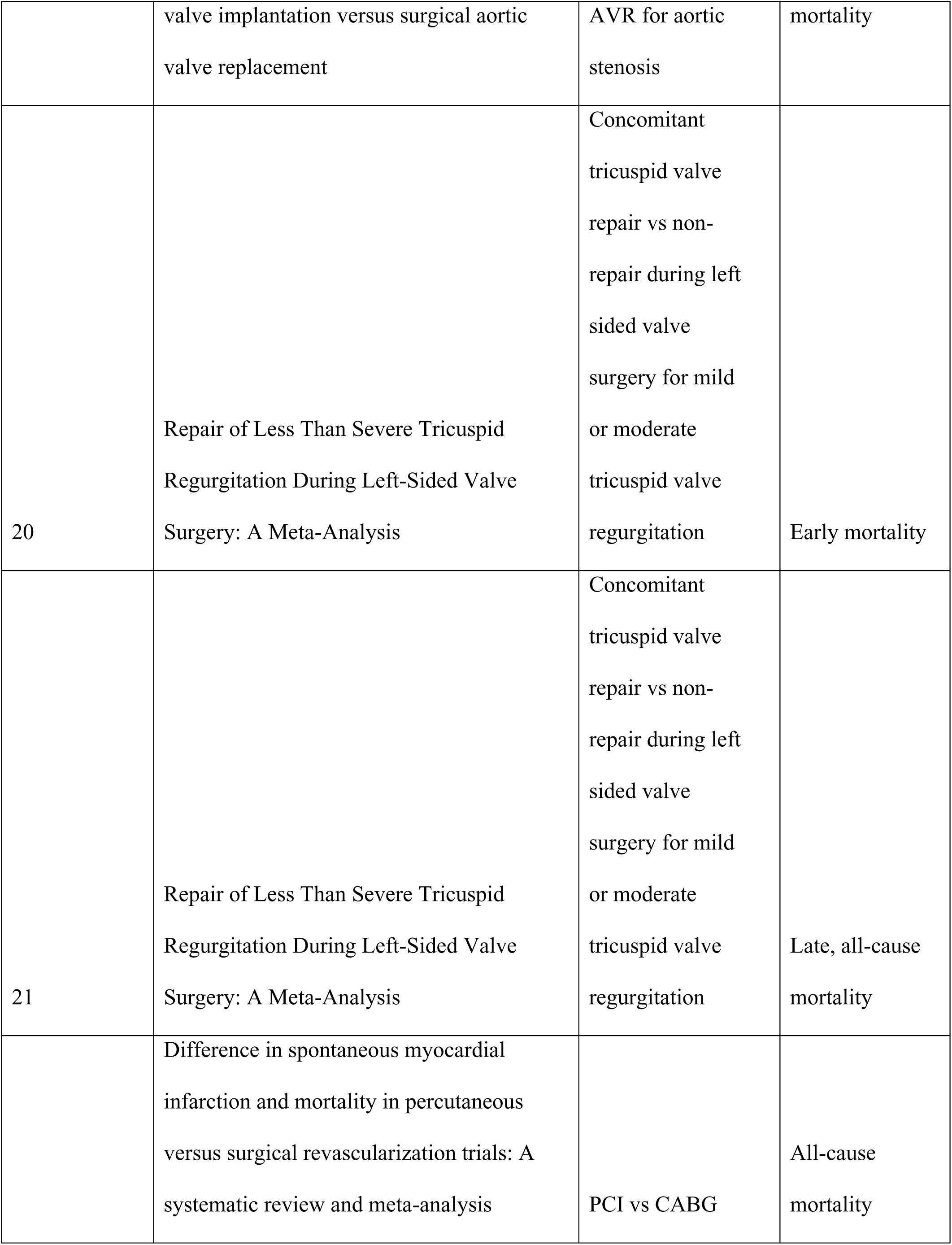
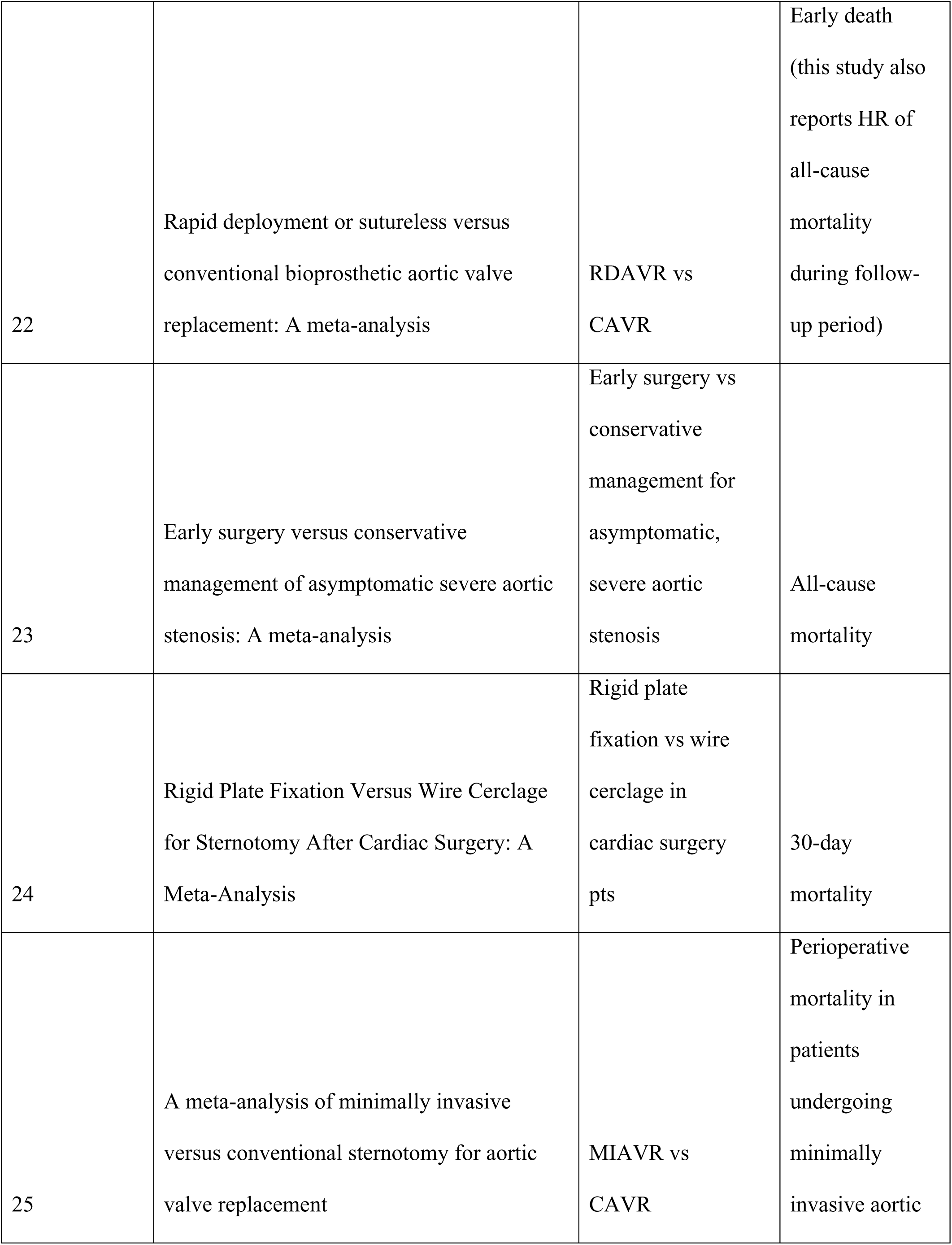
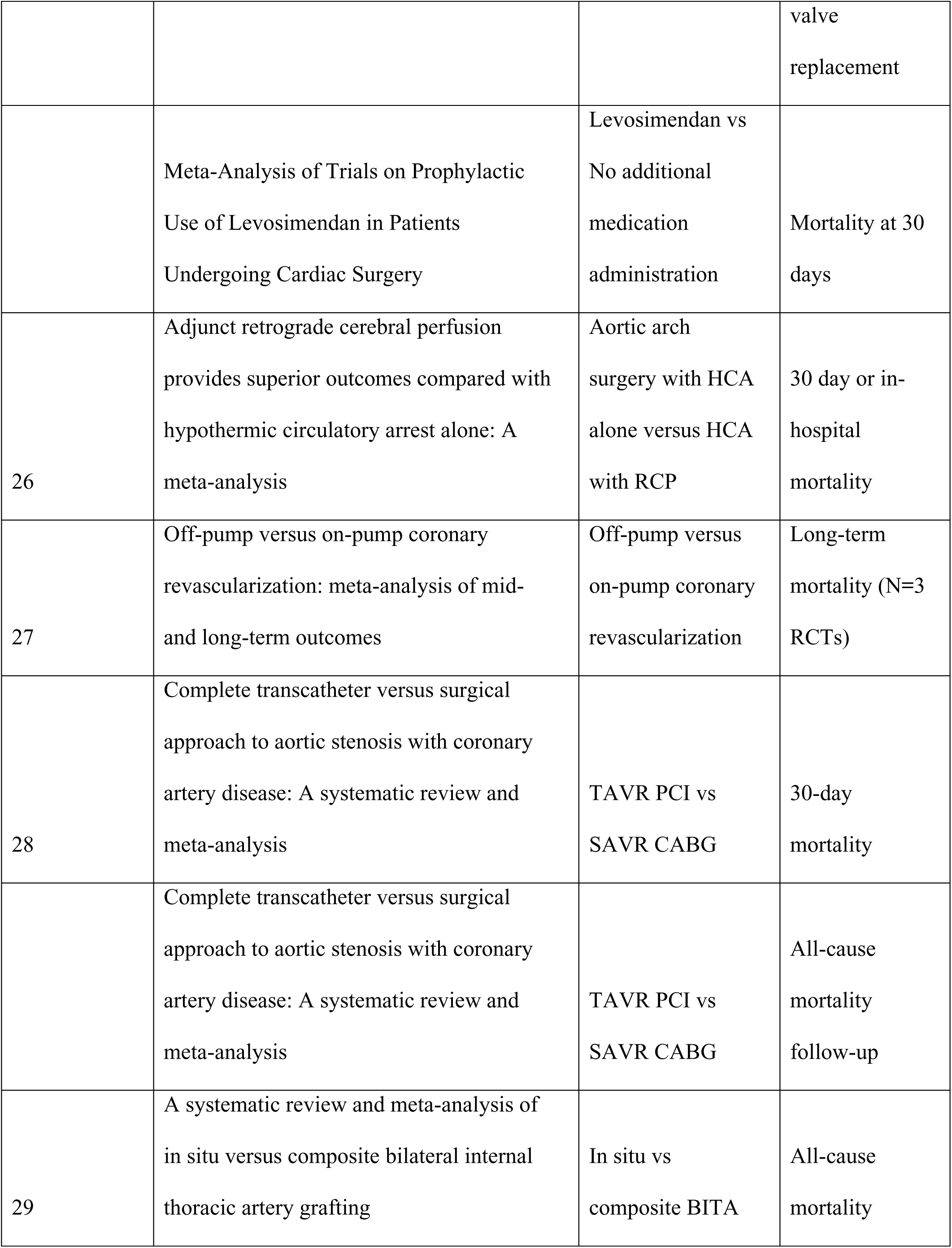
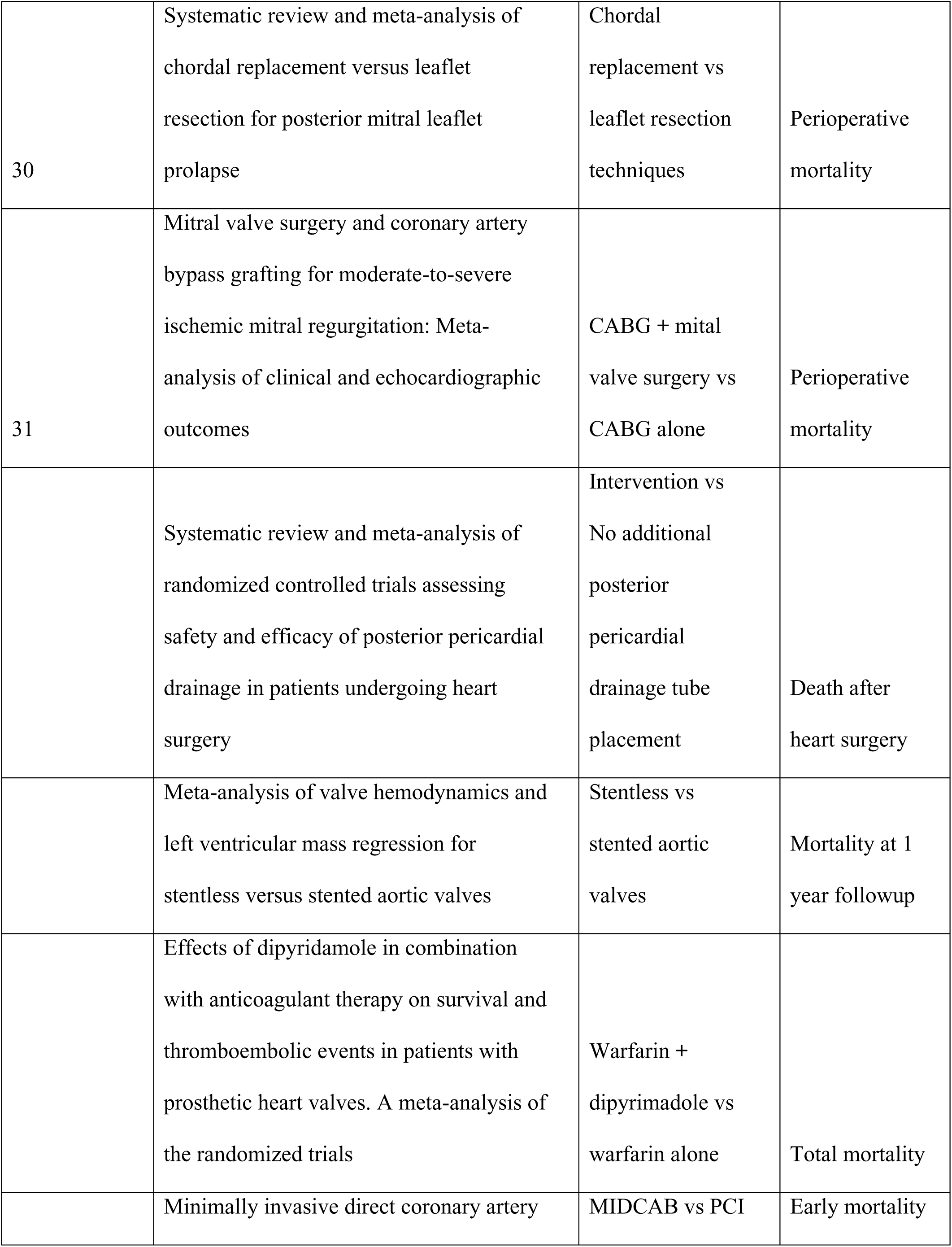
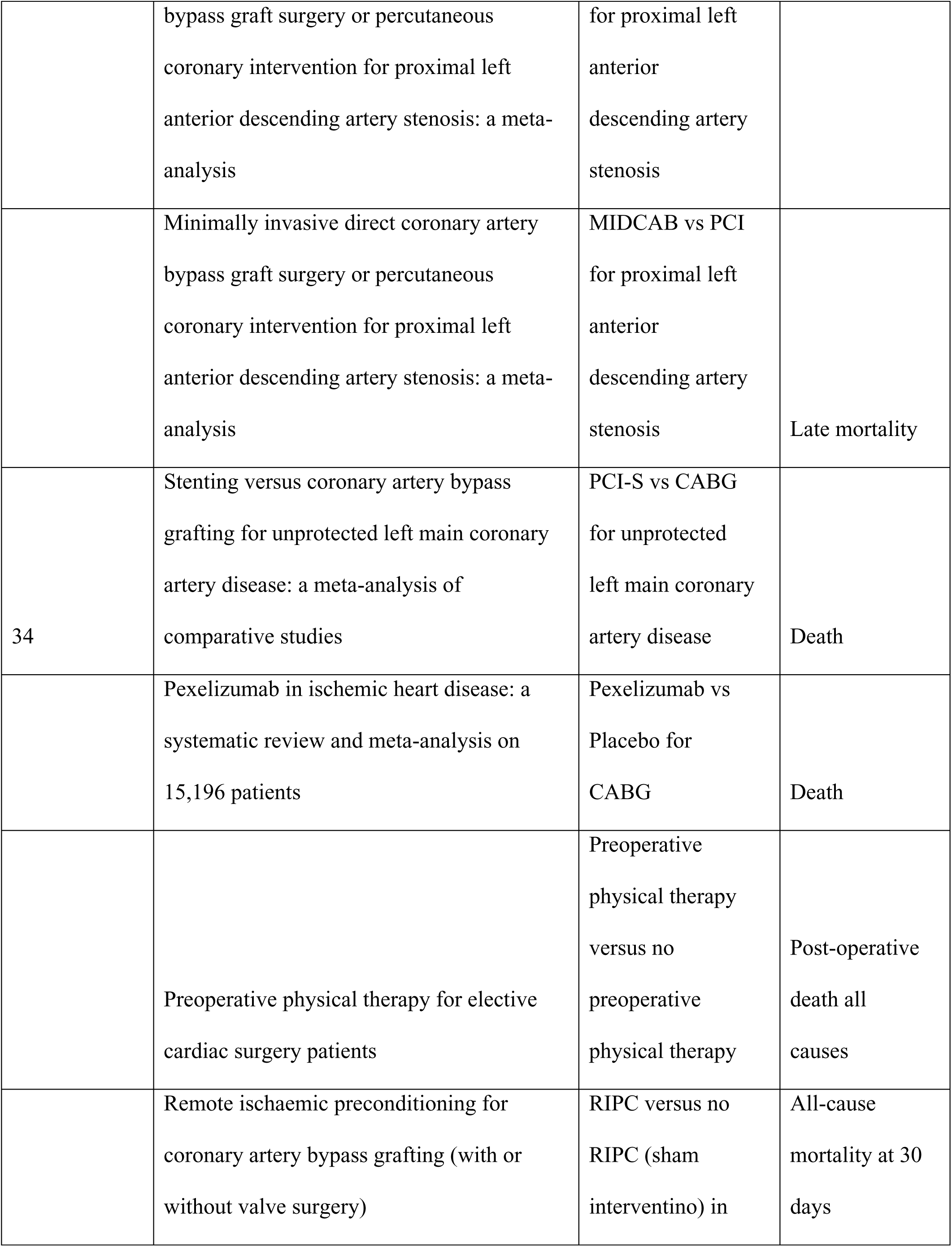
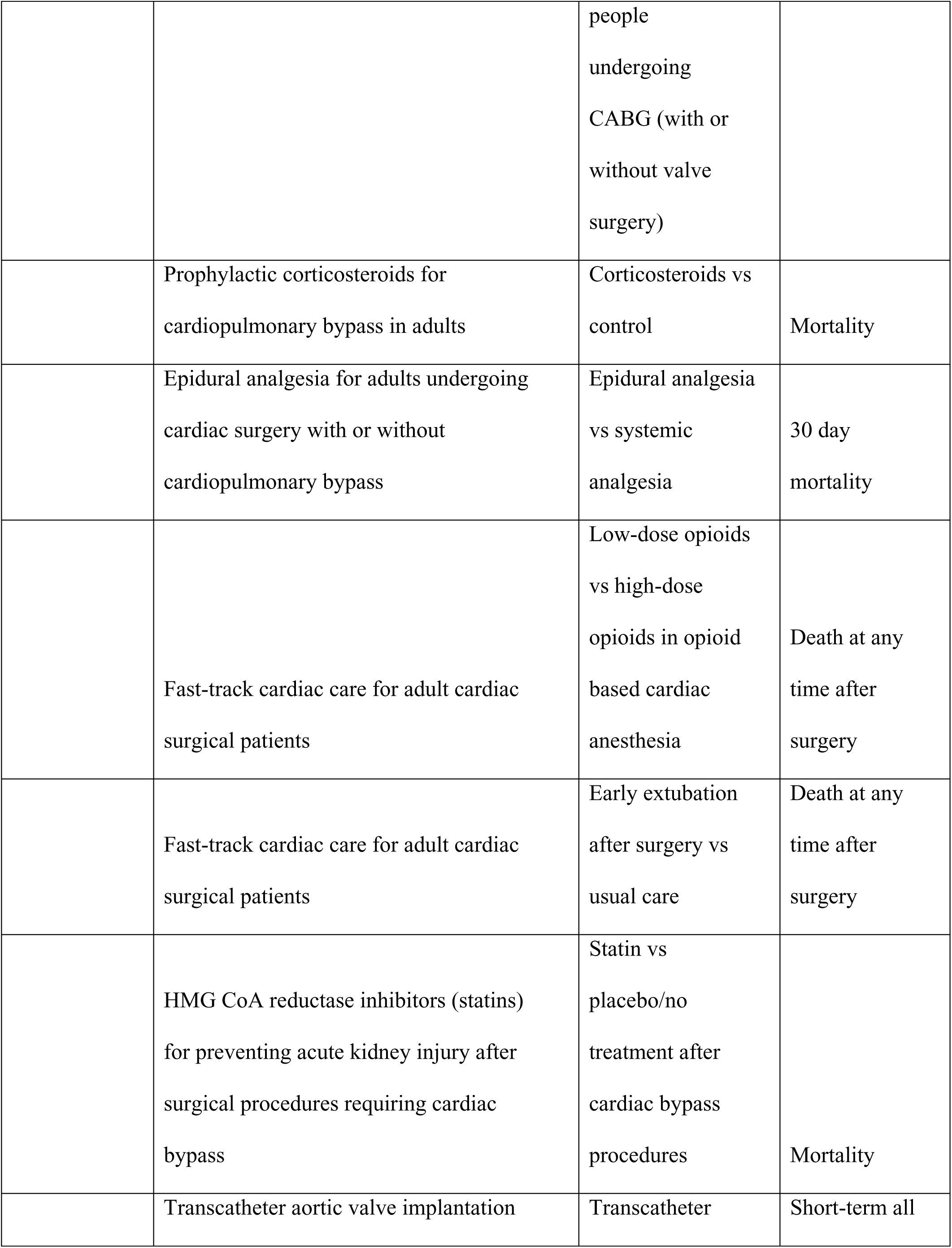
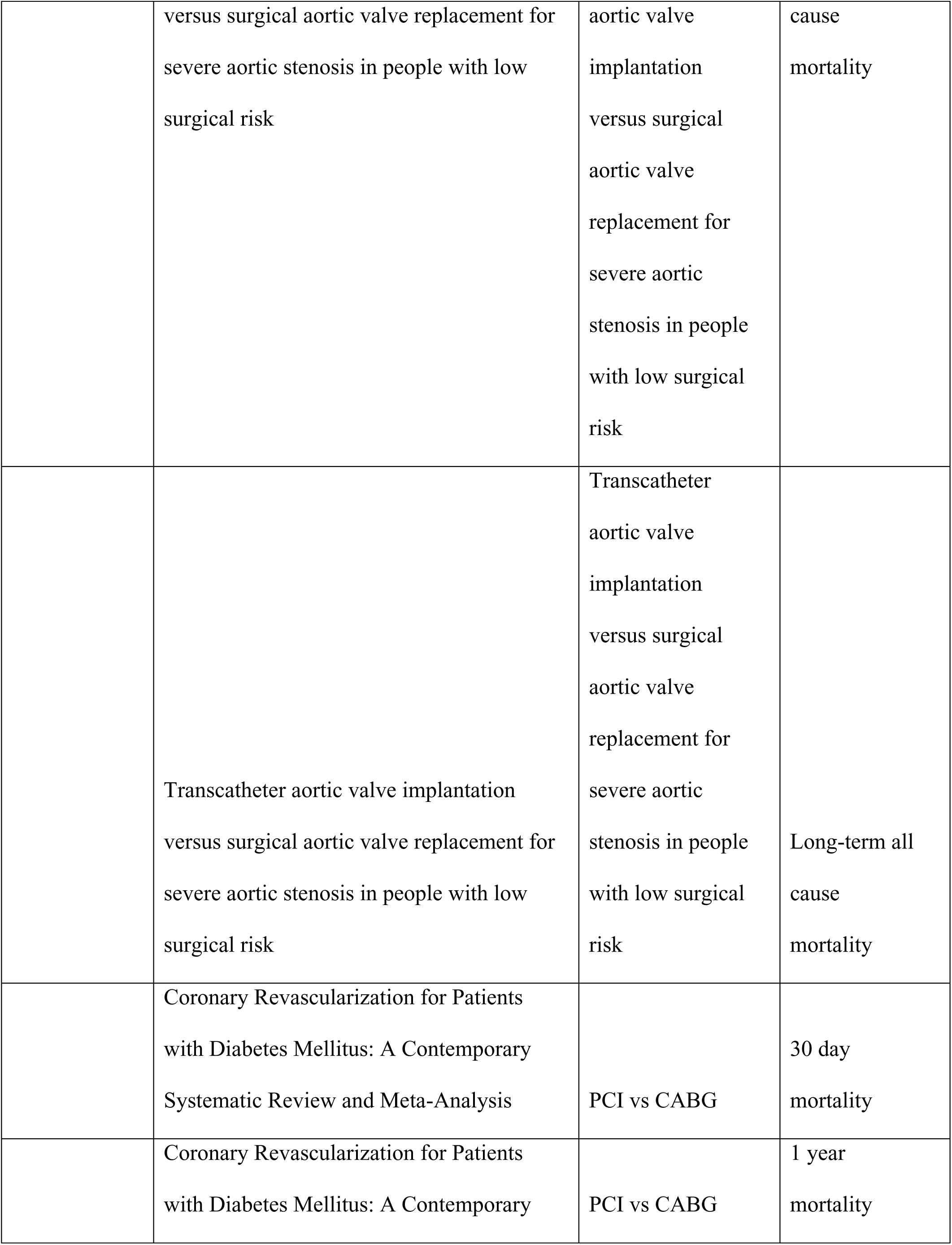
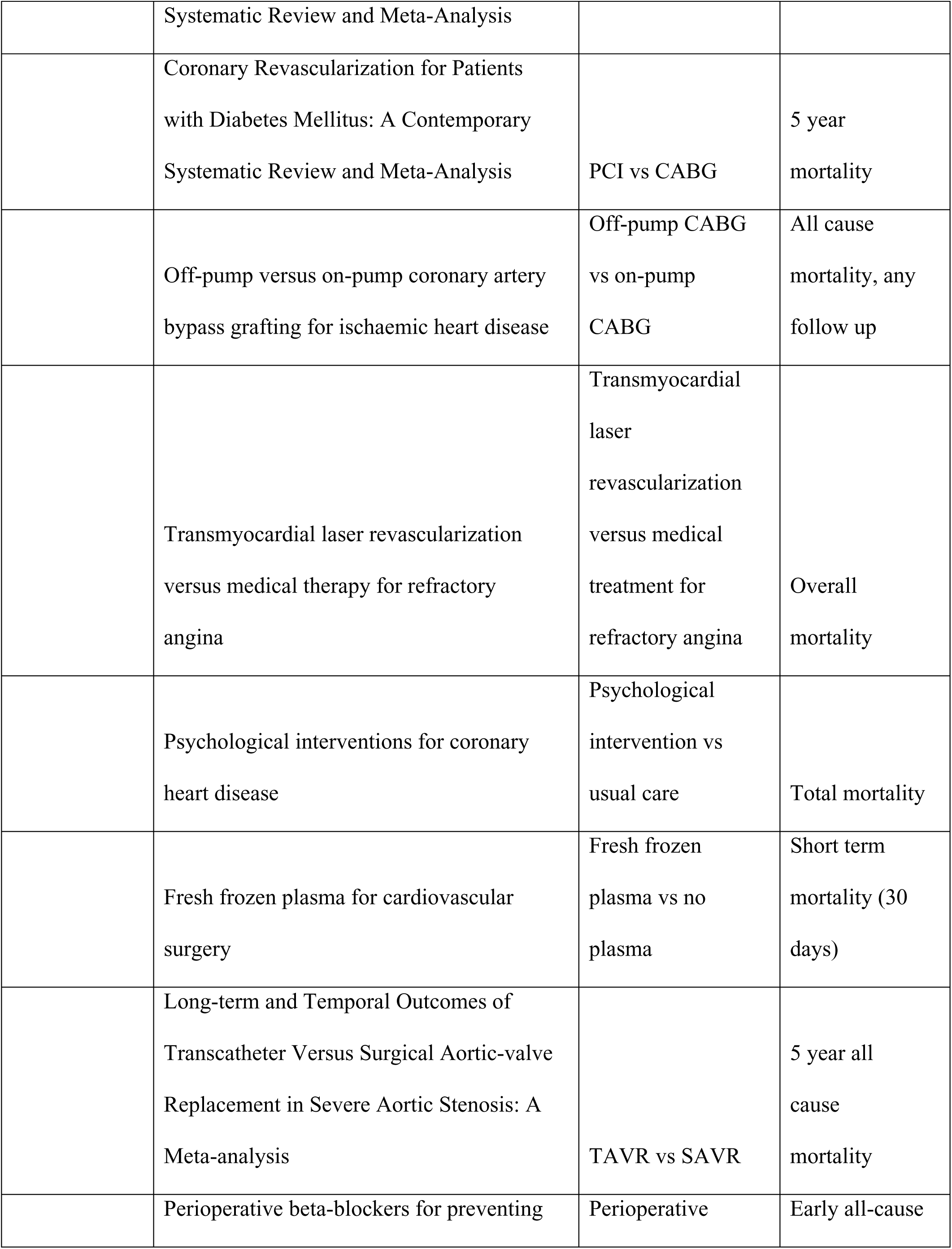
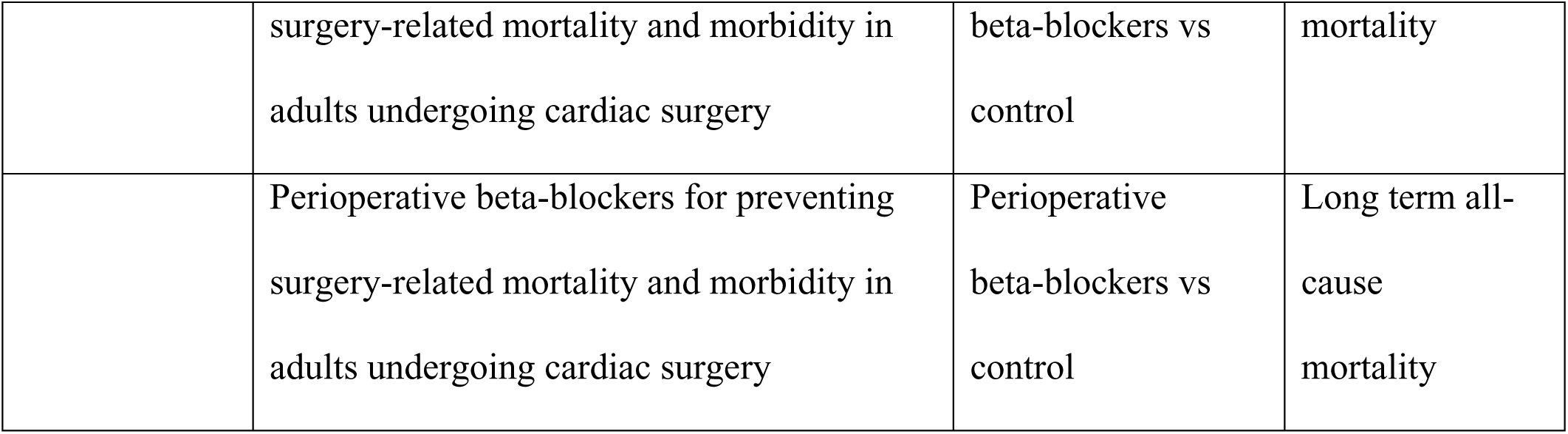
73 comparisons (comparison-outcome pairs) across 61 SRMAs. *Abbreviations*: ECMO: Extra Corporeal Membrane Oxygenation. PCI: Percutaneous Coronary Intervention. CABG: Coronary Artery Bypass Grafting. ARDS: Acute Respiratory Distress Syndrome. DAPT: Dual Anti-Platelet Therapy. SAPT: Single Anti-Platelet Therapy. TAVR: Transcatheter Aortic Valve Replacement. AVR: Aortic Valve Replacement. IABP: Intra-Aortic Balloon Pump. TVA: Tricuspid Valve Annuloplasty. OPCAB: Off Pump Coronary Artery Bypass. SAVR: Surgical Aortic Valve Replacement (same as AVR). MIDCAB: Minimally Invasive Direct Coronary Artery Bypass.

## Supplementary Results: Sensitivity analyses

In the 34 meta-analyses containing both RCTs and observational studies, the earliest published study was an RCT in 8 (24%). In the 26 meta-analyses where observational studies were published first, the median years until an RCT was published was 3 (IQR 1 to 6). Across these meta-analyses, comparing only the observational studies published before the first RCT appeared, the summary of the relative summary effects was 0.93 (95% CI, 0.67-1.28), p=0.64, I^2^=4% (0 to 34%).

Of 6 meta-analyses where the observational effect was statistically significant (p<0.005), only 1 also showed the RCT effect to be statistically significant. Of 2 meta-analyses where the RCT effect was statistically significant (p<0.005), 1 also showed a significant observational effect.

Of the 824 studies, 416 (50%) were type A comparisons (comparing a newer less invasive method versus an older more invasive method looking at non-inferiority). In sensitivity analysis flipping the coining of type A comparisons, the median observational summary effect across the 34 comparisons with both RCTs and observational studies was 0.89 (IQR, 0.1 to 1.16) versus an RCT effect of 0.86 (IQR, 0.59 to 1.03) (p=0.47). The median relative summary effect was 1.16 (IQR, 0.80 to 1.63) and the summary of the relative summary effects 1.03 (0.82 to 1.31, p=0.79).

## Notes

### Competing Interest Statement

The authors have declared no competing interest.

### Funding Statement

No funding has been received by any of the authors for this study.

### Author Declarations

No IRB required since this is an umbrella review of previously published meta analyses.

## References

(1) Cutler EC, Levine SA. Cardiotomy and valvulotomy for mitral stenosis. Boston Med Surg J 1923;188:1023–7.

(2) Gibbon JH Jr. Application of a mechanical heart and lung apparatus to cardiac surgery. Minn Med 1954;37(3):171–185.

(3) Coronary Artery Surgery Study (CASS): a randomized trial of coronary artery bypass surgery. Survival data. Circulation 1983;68(5):939–950.

(4) Carabello BA. Clinical practice. Aortic stenosis. N Engl J Med 2002:346–677.

(5) David TE. Aortic valve repair and aortic valve-sparing operations J Thorac Cardiovasc Surg 2015;149:9.

(6) Carpentier A. Cardiac valve surgery: the “grench correction.” J Thorac Cardiovasc Surg 1983;86:323.

(7) Dein JR, Frist WH, Stinson EF, et al: Primary cardiac neoplastms: early and late results of surgical treatment in 42 patients. J Thorac Cardiovasc Surg 1987;93:502.

(8) Bossert Torsten B, Gummert JF, Battellini, et al. Surgical experience with 77 primary cardiac tumors. Interact Cardiovasc Thorac Surg 2005;4:311–315.

(9) Barkun JS, Aronson JK, Feldman LS, Maddern GJ, Strasberg SM, Balliol Collaboration, et al. Evaluation and stages of surgical innovations. Lancet. 2009;374:1089–96.

(10) Gelijns AC, Ascheim DD, Parides MK, Kent KC, Moskowitz AJ. Randomized trials in surgery. Surgery. 2009;145:581–7.

(11) Ergina PL, Cook JA, Blazeby JM, Boutron I, Clavien P-A, Reeves BC, et al. Challenges in evaluating surgical innovation. Lancet. 2009;374:1097–104.

(12) Anglemyer A, Horvath HT, Bero L. Healthcare outcomes assessed with observational study designs compared with those assessed in randomized trials. Cochrane Database Syst Rev. 2014 Apr 29;2014(4):MR000034.

(13) Hemkens LG, Contopoulos-Ioannidis DG, Ioannidis JP. Agreement of treatment effects for mortality from routinely collected data and subsequent randomized trials: meta-epidemiological survey. BMJ. 2016 Feb 8;352:i493.

(14) Ioannidis JP, Haidich AB, Pappa M, Pantazis N, Kokori SI, Tektonidou MG, Contopoulos-Ioannidis DG, Lau J. Comparison of evidence of treatment effects in randomized and nonrandomized studies. JAMA. 2001 Aug 15;286(7):821–30.

(15) Lozada-Martinez ID, Ealo-Cardona CI, Marrugo-Ortiz AC, Picón-Jaimes YA, Cabrera-Vargas LF, Narvaez-Rojas AR. Meta-research studies in surgery: a field that should be encouraged to assess and improve the quality of surgical evidence. Int J Surg. 2023 Jun 1;109(6):1823–1824.

(16) IntHout J, Ioannidis JP, Borm GF. The Hartung-Knapp-Sidik-Jonkman method for random effects meta-analysis is straightforward and considerably outperforms the standard DerSimonian-Laird method. BMC Med Res Methodol. 2014 Feb 18;14:25.

(17) Page MJ, McKenzie JE, Bossuyt PM, Boutron I, Hoffmann TC, Mulrow CD, Shamseer L, Tetzlaff JM, Akl EA, Brennan SE, Chou R, Glanville J, Grimshaw JM, Hróbjartsson A, Lalu MM, Li T, Loder EW, Mayo-Wilson E, McDonald S, McGuinness LA, Stewart LA, Thomas J, Tricco AC, Welch VA, Whiting P, Moher D. The PRISMA 2020 statement: An updated guideline for reporting systematic reviews. Int J Surg. 2021 Apr;88:105906.

(18) Sedgwick P. Meta-analyses: what is heterogeneity? BMJ. 2015 Mar 16;350:h1435.

(19) von Hippel PT. The heterogeneity statistic I(2) can be biased in small meta-analyses. BMC Med Res Methodol. 2015 Apr 14;15:35.

(20) Balduzzi S, Rücker G, Schwarzer G. How to perform a meta-analysis with R: a practical tutorial. Evid Based Ment Health. 2019 Nov;22(4):153–160.

(21) de Winter JCF, Gosling SD, Potter J. Comparing the Pearson and Spearman correlation coefficients across distributions and sample sizes: A tutorial using simulations and empirical data. Psychological Methods 2016;21(3):273–290.

(22) Kjaergard LL, Villumsen J, Gluud C. Reported methodologic quality and discrepancies between large and small randomized trials in meta-analyses. Ann Intern Med. 2001 Dec 4;135(11):982–9.

(23) Benjamin DJ, Berger JO, Johannesson M, Nosek BA, Wagenmakers EJ, Berk R, et al. Redefine statistical significance. Nat Hum Behav. 2018 Jan;2(1):6–10.

(24) Gerstein HC, McMurray J, Holman RR. Real-world studies no substitute for RCTs in establishing efficacy. Lancet. 2019 Jan 19;393(10168):210–211.

(25) Djulbegovic B, Glasziou P, Chalmers I. The importance of randomised vs non randomised trials. Lancet. 2019 Aug 24;394(10199):634–635.

(26) McCulloch P, Altman DG, Campbell WB, Flum DR, Glasziou P, Marshall JC, et al. No surgical innovation without evaluation: the IDEAL recommendations. Lancet. 2009;374:1105–12.

(27) Ioannidis JPA. Options for publishing research without any P-values. Eur Heart J. 2019 Aug 14;40(31):2555–2556.

(28) Amrhein V, Greenland S, McShane B. Scientists rise up against statistical significance. Nature. 2019 Mar;567(7748):305–307.

(29) Ioannidis JPA. Retiring statistical significance would give bias a free pass. Nature. 2019 Mar;567(7749):461.

(30) Koletsi D, Solmi M, Pandis N, Fleming PS, Correll CU, Ioannidis JPA. Most recommended medical interventions reach P < 0.005 for their primary outcomes in meta-analyses. Int J Epidemiol. 2020 Jun 1;49(3):885–893.

(31) Kolaski K, Logan LR, Ioannidis JPA. Guidance to best tools and practices for systematic reviews. JBI Evid Synth. 2023;21(9):1699–1731.

